# The burden of Intestinal helminthic infection and Anaemia among pregnant women in Kabwe, Zambia; a Cross-Sectional study

**DOI:** 10.1101/2024.11.26.24317960

**Authors:** Mellah Mulenga Kasoma, Amon Siame, Westone Hamwata, Nzooma M. Shimaponda-Mataa

## Abstract

**Background:** Zambia is a developing country where intestinal helminthic infections are a serious public health concern. In sub-Saharan Africa alone it is estimated that one-third of pregnant women are infected with soil-transmitted helminthes which can lead to anaemia in pregnancy. The burden of anaemia, associated with helminths, is an important contributor to maternal and fetal mortality. This study aimed to determine the prevalence of intestinal helminthic infection and anaemia among pregnant women attending Antenatal Care (ANC) in Kabwe, Zambia.

**Methodology:** A health facility-based cross-sectional study was conducted among 442 pregnant women from November,2021, to February, 2022, in Kabwe, Zambia. Data were collected using pre-tested questionnaires from three health facilities. Blood and stool specimens were processed using standard operating procedures. Cleaned and coded data were entered and analyzed using SPSS version 28. Multivariate binary logistic regression analysis was used to determine the association of predictors and response variables at P <0.05. Adjusted odds ratio with 95% CI was used to show the strength of association between predictors and outcome variables.

**Results:** A total of 442 pregnant women were enrolled in this study with a median age of 23 years (range 15–41 years). The total prevalence of intestinal helminths was 24.7% (75/442) with the predominance of Hookworm (13.3%) followed by *Ascaris lumbricoides* (2.3%). Eating raw vegetables (AOR= 19.67, 95% CI: 1.20 to 321.69), drinking water from the well (AOR=2.45, 95% CI: 1.17 to 5.17) and Walking barefoot (AOR=0.25, 95% CI: 0.10 to 0.60) were significant predictors of intestinal helminthic infection. The overall prevalence of anaemia was 64.9% (281/442).

**Conclusion:** The prevalence of intestinal helminths and anaemia was significantly high in this study. Eating raw vegetables, walking barefoot and drinking water from the well were identified as significant contributors to intestinal helminthic infection and anaemia among pregnant women. Therefore, public health measures and intensive antenatal care services are vital to promoting safe pregnancy.

**Author Summary:** Many pregnant women in underdeveloped nations suffer from intestinal helminthic infections and anaemia, which raises their risk of dying during pregnancy and giving birth to underweight babies, who are also more likely to die. Although it has long been known that human worm infection is one of the main causes of anaemia in underdeveloped areas, knowledge of the advantages of treating worm infection during pregnancy has lagged behind that of other primary causes of maternal anaemia. Low coverage of anthelmintic treatment in maternal health programmes in many countries including ours has been the result. After conducting the study Intestinal worms and anaemia were diagnosed in 24.7% and 64.9% of pregnant women in Central Zambia who provided stool and blood samples. In these women, intestinal worm infections caused a modest decrease in haemoglobin (Hb) levels. We observed that increasing worm infection intensity is associated with lower haemoglobin levels in pregnant women. We also estimate that between a quarter and a third of pregnant women in sub-Saharan Africa are infected with worms and at risk of preventable anaemia. Nonetheless, every intervention study that was found demonstrated the advantages of deworming for the health of the mother or child, and we contend that more should be done to ensure that pregnant women receive anthelmintic therapy.

## Introduction

Intestinal parasitic infection (IPI) is a serious public health problem throughout the world, particularly in developing countries with an estimated 3.8 billion people infected, 720 million cases, and an estimated 135,000 deaths annually. They are among the Neglected Tropical Diseases (NTDs) of poverty [1]. Soil-transmitted helminths (STH) also known as intestinal helminths is a term referring to a group of parasitic diseases caused by nematode worms that are transmitted to humans by faecally contaminated soil and packages three intestinal parasites *Trichuris trichuira*, hookworm, and Ascaris lumbricoides [2]. Zambia is a developing country where intestinal helminths are a major public health problem. Previous studies carried out in Zambia in children revealed a high prevalence of intestinal infection. The burden of intestinal parasites, particularly the STHs, is often very high in school children and pregnant women as high-risk populations [4,5]. Anaemia is a medical condition in which there is less than the normal haemoglobin (Hb) level in the body, which decreases the oxygen-carrying capacity of red blood cells to tissues. Anaemia is more prevalent in developing countries arising from poor nutritional status and the high prevalence of parasitic infection. Women’s health is significantly impacted by anaemia, particularly in developing countries. Anaemia is a major factor in women’s health, especially during pregnancy for when severe, it significantly contributes to maternal mortality [6]. Anaemia during pregnancy is defined as a haemoglobin concentration of less than 11 grams per deciliter (g/dL)[7].

Although there are several studies conducted in Zambia that have reported the magnitude of intestinal parasitic infections. These studies focus on children as being at high risk. There is a paucity of published data on intestinal helminths and anaemia in the study area particularly in pregnant women as an additional at-risk population. Land cultivation is the major economic activity in Kabwe district which plays a crucial role in the prevalence and transmission of STHs particularly in communities with poor sanitation. Additionally, the humid climate of the district is also suitable for the development of intestinal helminths. Furthermore, the availability of street-vending fruits and the habit of wearing open shoes may also increase the prevalence of intestinal helminths in Kabwe district of Zambia compared to other countries. The study’s results may help concerned stakeholders take action to prevent intestinal helminths and anaemia in pregnant women. This study, therefore, aimed to determine the prevalence of intestinal helminths and associated anaemia among pregnant women in Kabwe district, Zambia. The aim was achieved.

## Methods

### Study Setting and Context

A health facility-based cross-sectional study was conducted from November 2021 to February 2022 in three randomly selected health facilities in Kabwe district, Zambia: Makululu Health Centre, Katondo Clinic, and Kasanda Health Centre.

### Study Population

Our study population were all pregnant women in the reproductive age 15 to 49 years old who were attending antenatal services at the selected health centres of Katondo, Kasanda and Makululu health centres in Kabwe District (14.4285° S, 28.4514° E). The World Health Organization (WHO) defines reproductive age as the period between 15 and 49 years. [8]. The study strictly excluded all the pregnant women who were taking anthelminthic/anti-protozoan drugs within 6 months before the study and those with confirmed acute and/or chronic diseases that caused anaemia.

### Sample Size Determination and Sampling Procedure

The sample size was estimated using a formula for survey sample size estimation taking 95% level of confidence.

The sample size was estimated using a formula for survey sample size estimation at 95% level of confidence.

Where; 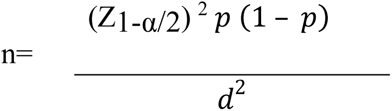

n = sample size, Z_1-α/2_= value of the standard normal distribution (1.96) corresponding to a significance level of 0.05 for a 2-sided test, d = margin of error (0.05), the prevalence (p= 0.41) assumption was based on a survey conducted in Ethiopia [9]

Sample size (n) = 372, adding by 19% refusal rate The final sample size was 442.

To attain the desired unbiased sample size, a systematic random sampling method was used to recruit the study participants. Every 3rd pregnant mother who visited ANC was consecutively enrolled until the planned sample size was achieved.

### Data Collection

Before the actual data collection, the questionnaire was pre-tested on 21 pregnant women (5% of the total sample size) who were attending ANC in an un-selected health facility, Pollen Health Centre. Pregnant women were informed about the objective of the study before data collection and semi-structured questionnaires were used to obtain data on socio-demographic, and environmental related factors, obstetric characteristics, and dietary habits. The questionnaire was developed in English and participants were interviewed using the local language; Bemba. Medical Laboratory professionals who can speak the local language were trained in data collection for this particular study to attain standardization, maximize interviewer reliability and minimize bias. The data collectors were regularly supervised by the investigators for proper data collection. All reagents used were checked for their expiry date and prepared according to the manufacturer’s instructions.

### Specimen Collection and Processing

#### Blood Collection

Blood was collected using micro cuvettes to measure the haemoglobin levels (Hb). Haemoglobin levels were measured on-site using HemoCue Hb 301 (HemoCue AB, Angelholm Sweden). Blood collection was done using a needle finger prick on the middle finger of the non-dominant hand the fingertips were first cleaned and disinfected with an antiseptic solution and allowed to dry before being lightly pressed by the phlebotomist using their thumb from the top of the knuckle towards the fingertips to stimulate blood flow. While pressing the finger lightly towards the fingertip, a lancet was used to puncture the fingertip. The first drop of blood was wiped. Then the finger was pressed lightly towards the fingertip until another drop of blood appeared. A microcuvette was filled in one step (approximately 10 microliter) of blood when the blood drop was large enough. The microcuvette was wiped from the outside to ensure that no specimen was drawn from the end, it was then placed in the cuvette holder and the measurement was started by gently sliding the cuvette holder to the measuring position. After about three seconds, the haemoglobin value was displayed and recorded after which the micro cuvette was removed and discarded. All the readings were done within 40 seconds of loading. Anemia was defined based on WHO criteria as; anemia: haemoglobin level <11 g/dL for pregnant women.

#### Stool Collection

Stool specimen was obtained from all patients selected for the study. A direct saline and iodine wet mount of each sample was used to detect intestinal parasites microscopically. The wet mounts were examined under a light microscope at 10*×* and 40*×.* A portion of each stool specimen was then taken and processed using formolether concentration technique following standard procedures. Briefly, 1 g of stool was placed in a clean conical centrifuge tube containing 7 ml 10% formol water using an applicator stick. The resulting suspension was filtered through a sieve into another conical tube. After adding 3–4 ml of diethyl ether to the formalin solution, the content was centrifuged at 3200 rpm for 1 min. The supernatant was discarded; a smear was prepared from the sediment and observed under a light microscope with a magnification of 100*×* and 400*×* [9]. Stool samples were recorded as positive with the presence of ova.

### Quality Control

One senior midwife who was working in the ANC department at each selected health facilities and two Health workers were recruited as research assistants who were under the whole supervision of the principal investigator. Two days of training were conducted for data collectors to aid in the clarity, consistency, and reliability of measurements. Collected data were checked for consistency and completeness daily and pre-analytical, analytical and post-analytical laboratory data quality were controlled accordingly. Quality control was performed on the analytical instrument (HemoCue) according to the established quality control guidelines and to minimize observer bias three different laboratory technologists independently examined the stool microscopy slides before result confirmation. All laboratory analyses were carried out using standard operating procedures.

### Data Analysis

The data was cleaned, coded and double-entered. Excel was used for data entry. The data was exported to SPSS version 28 (SPSS INC, Chicago, IL, USA) software for analysis. Missing data were managed by observing cross-tabulation results and percentages. Firstly, all numerical variables including maternal age and haemoglobin concentration were tested for normality using the Shapiro-Wilkt test. Pearson’s Chi-square test of association was performed to compare the haemoglobin levels with sociodemographic, lifestyle and parasitic infections. Additionally, a non-parametric test, Mann-Whitney U-Test was used to explore the correlation between the distribution of haemoglobin levels and the different parasitic infections. Lastly, Binary logistic regression models were used to determine the association between predictors and outcome variables. Bivariate analysis was carried out for each independent variable to identify the presence of association with the response variable at P < 0.25 Multivariate logistic regression was used to control the possible confounding factors at P < 0.05. Adjusted odds ratio with 95% confidence (AOR, 95%CI) was used to infer the results and all data analyses were done according to standard protocols [5]

### Ethical Considerations and Permissions

This study involved direct contact with humans and also had potential to invade participant privacy given that stool collection is confidential and uncomfortable. As such, the study was approved by the Research Ethics Committee of The University of Zambia. Protocol ID 202112030113. Then an official letter of cooperation was written to the Kabwe District Office from The University of Zambia and permission was obtained. All participants were old enough to provide informed consent in line with local laws. A verbal informed consent process was conducted for participants who could not read and write. Participants’ right during the interview for either not to participate or to withdraw at any stage of the interview was guaranteed. All the information obtained from the study participants was coded to maintain confidentiality and positive results were immediately communicated with clinicians for appropriate intervention.

## Results

### Socio-demographic Characteristics of Study Participants

A total of 442 pregnant women participated in this study. The median age of the women was 23 years (± 8) years and the majority (57.7%; 255/442) fell within 20-29 years. The majority of the women (91%; 382/442) had secondary school education level training. The majority of the women (63%; 279/442) in this study were single (Table 1)

**Table 1:**
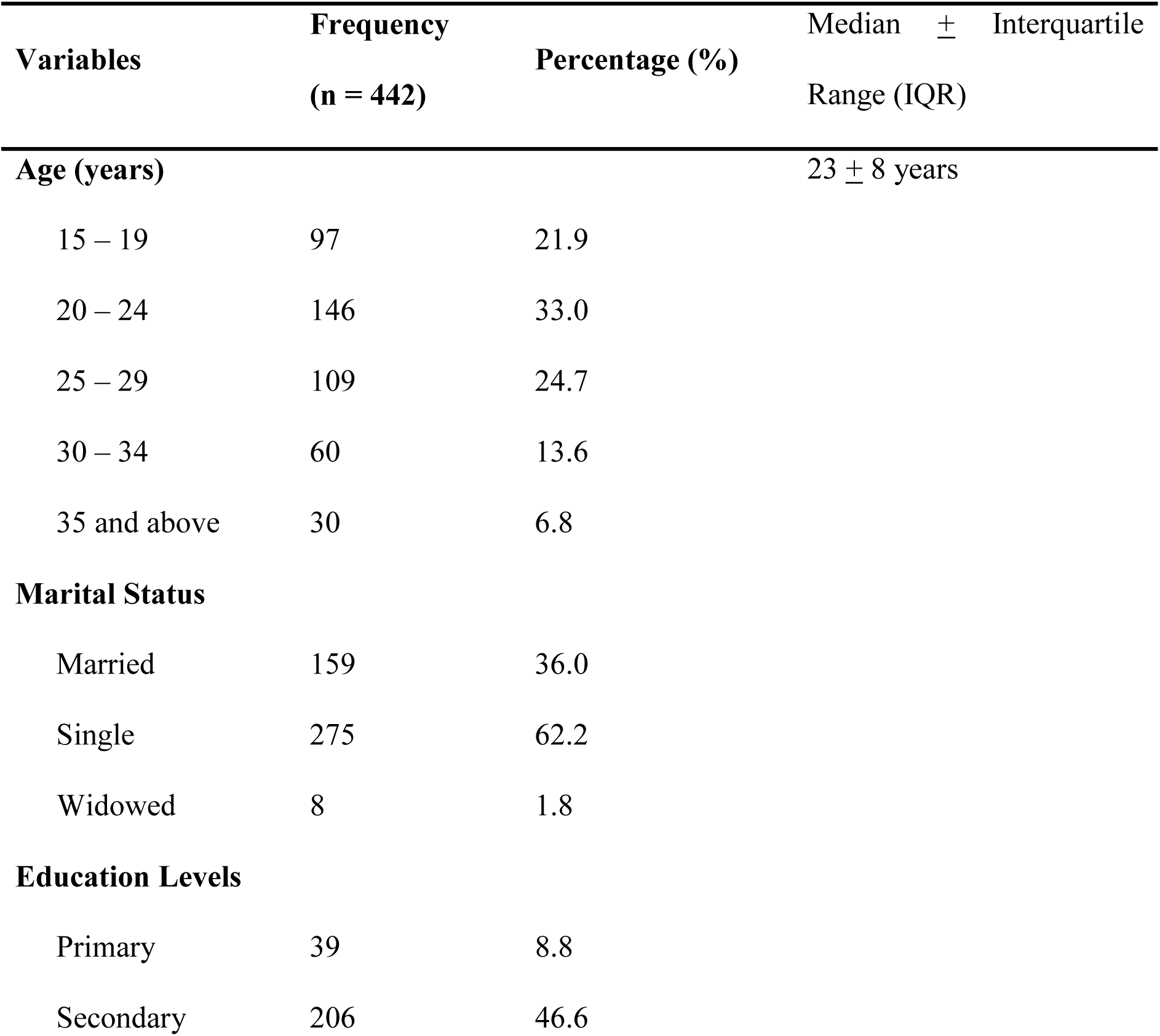
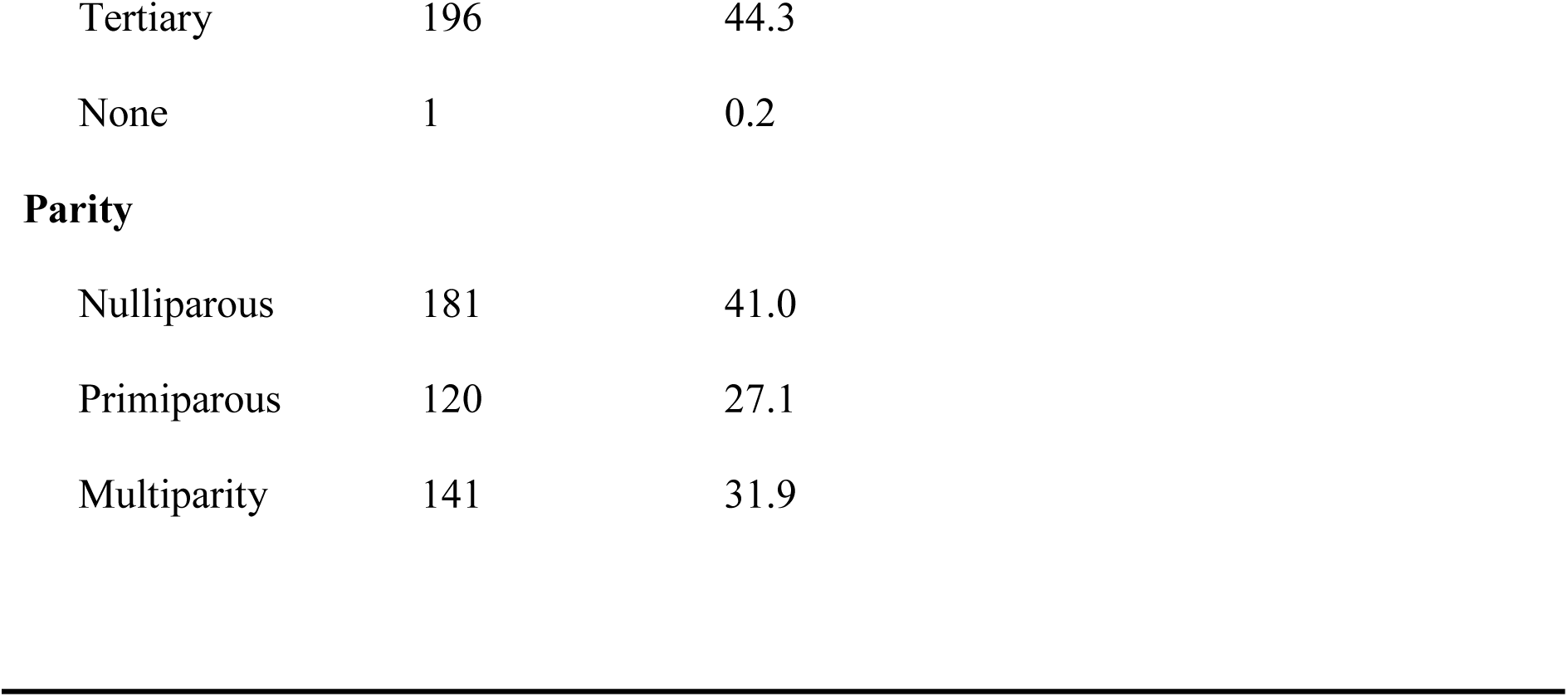
Sociodemographic Characteristics of Pregnant Women attending ANC in Kabwe, Zambia from November 2021 to February 2022.

| Variables | Frequency<br>(n = 442) | Percentage (%) | Median ± Interquartile<br>Range (IQR) |
| --- | --- | --- | --- |
| **Age (years)** | | | 23 ± 8 years |
| 15 – 19 | 97 | 21.9 |  |
| 20 – 24 | 146 | 33.0 |  |
| 25 – 29 | 109 | 24.7 |  |
| 30 – 34 | 60 | 13.6 |  |
| 35 and above | 30 | 6.8 |  |
| **Marital Status** |  |  |  |
| Married | 159 | 36.0 |  |
| Single | 275 | 62.2 |  |
| Widowed | 8 | 1.8 |  |
| **Education Levels** |  |  |  |
| Primary | 39 | 8.8 |  |
| Secondary | 206 | 46.6 |  |

|  |  |  |
| --- | --- | --- |
| Tertiary | 196 | 44.3 |
| None | 1 | 0.2 |

**Parity**
|  |  |  |
| --- | --- | --- |
| Nulliparous | 181 | 41.0 |
| Primiparous | 120 | 27.1 |
| Multiparity | 141 | 31.9 |

### Lifestyle and Behavioural Characteristics of Study Participants

Almost half of the women, 36.7% (162/442) walked barefoot when at home. The majority of the women, 84.8% (375/442) ate salads, a significant proportion of women, 93.9 % (415/442), washed their fruits before eating. Regarding the source of drinking water, the majority of the women, 79.2 % (35/442), rely on tap water, while 20.8% use well water. The study also found that a significant proportion of women, 64% (283/442) and 25.8 % (114/442) engage in gardening and farming activities, respectively. (Table 2)

**Table 2:** Lifestyle and Behavioural Characteristics of pregnant women attending ANC in Kabwe, Zambia from November 2021 to February 2022.

| **Variables** | **Frequency (n = 442)** | **Percentage (%)** |
| --- | --- | --- |
| **Eat Salads (raw vegetables)** |  |  |
| No | 64 | 14.5 |
| Yes | 375 | 84.8 |
| Sometimes | 3 | 0.7 |
| **Wash fruits before eating.** |  |  |
| No | 1 | 0.2 |
| Yes | 415 | 93.9 |
| Sometimes | 26 | 5.9 |
| **Walks Barefooted** |  |  |
| No | 162 | 36.7 |
| Yes | 162 | 36.7 |
| Sometimes | 118 | 26.6 |
| **Source of Drinking Water** |  |  |
| Tap water | 350 | 79.2 |
| Well water | 92 | 20.8 |
| **Does Gardening or Farming?** |  |  |
| No | 45 | 10.2 |
| Gardening only | 283 | 64.0 |
| Farming only | 114 | 25.8 |

### Deworming status of study participants

More than half 87.3% (386/442} of the respondents had never received deworming tablets during the antenatal visits. The study found that a significant proportion of participants had not been dewormed during the pregnancy or 6 months before the study. (Table 3)

**Table 3:** deworming status of pregnant women attending ANC in Kabwe, Zambia from November 2021 to February 2022.

| Variables | Frequency (n = 442) | Percentage (%) |
| --- | --- | --- |
| **Deworming status during pregnancy** |  |  |
| Never dewormed | 386 | 87.3 |
| Yes, but more than 6 months ago | 56 | 12.7 |

### Prevalence of Intestinal Helminth Infection in study participants

The total prevalence of intestinal helminths in this study was 17% (72/442). The results in Fig 1 show that the majority of the 442 study participants (83%) did not have any intestinal helminths. On the other hand, a significant proportion of the participants were infected with one or more species of intestinal helminths. The predominant intestinal helminth was hookworm which accounted for 13.3% (59/442) followed by *Ascaris lumbricoides* which was found in 2.3% of the participants. The combination of hookworms and *A. lumbricoides* was found in 0.7% of the participants, while the combination of hookworms and *S. stercoralis* was found in 0.5% of the participants. The least common infections were *Trichuris trichura* and hookworms, *S. stercoralis*, and *A. lumbricoides*, which were found in 0.2%, 0.5%, and 0.7% respectively.

**Figure 1:**
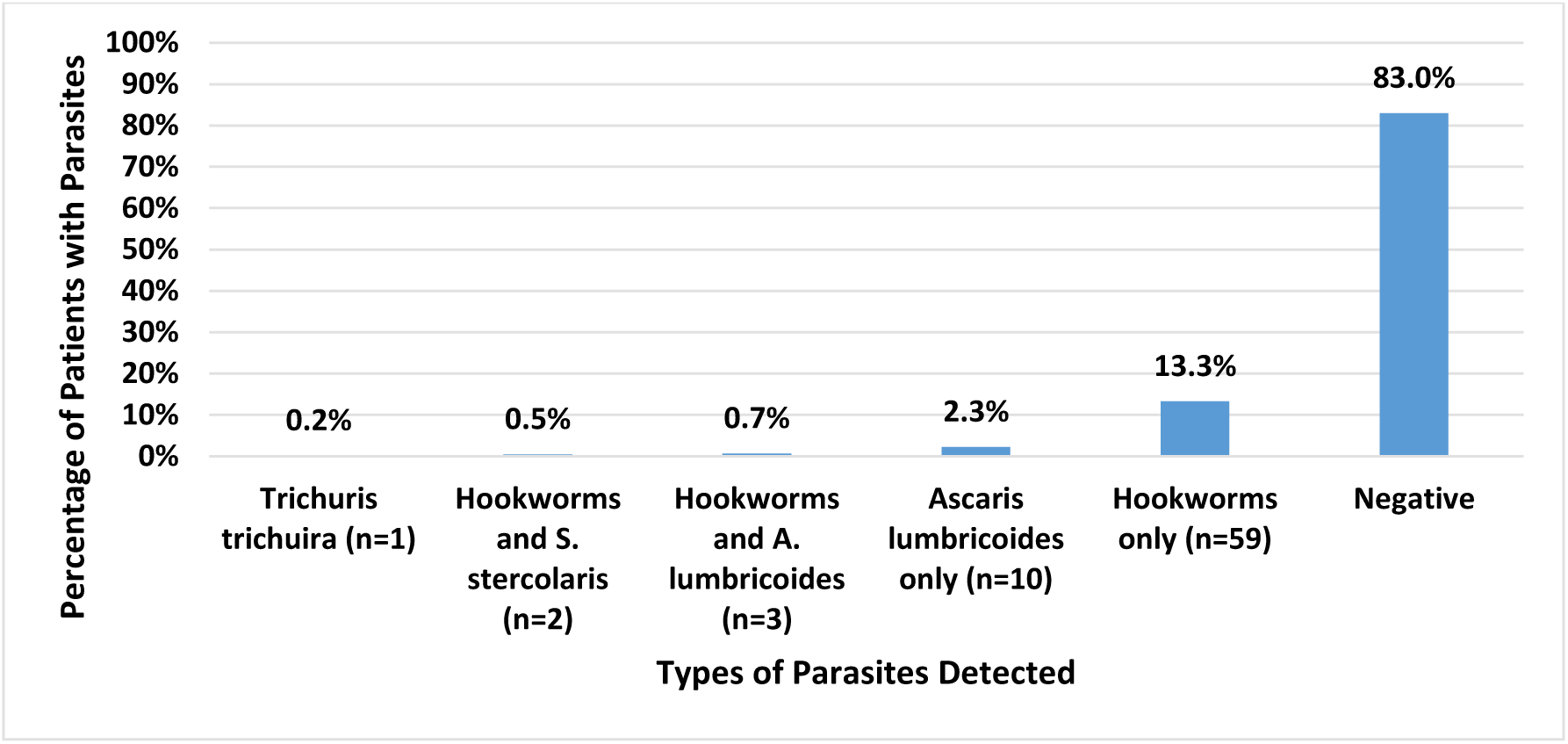
Bar Chart Showing the Distribution of Parasites in pregnant women attending ANC in Kabwe, Zambia from November 2021 to February 2022

### Haemoglobin levels and Anaemia status in the study participants

The overall prevalence of anaemia in this study was 35.1% (155/442) with the median haemoglobin level of the pregnant women being 11.7 g/dL+ 2.1 g/dL (Table 5)

**Table 4:** Haemoglobin Levels of pregnant women attending ANC in Kabwe, Zambia from November 2021 to February 2022.

| Variables | Frequency<br>(n = 442) | Percentage<br>(%) | Median ± Interquartile Range<br>(IQR) |
| --- | --- | --- | --- |
| **Haemoglobin Level** |  |  |  |
| **(g/dL)** |  |  |  |
| $\geq 11 \text{ g/dl}$ (Normal) | 287 | 64.9 | $11.7 \pm 2.1 \text{ g/Dl}$ |
| $< 11 \text{ g/dl}$ (Anaemic) | 155 | 35.1 | |

**Table 5:**
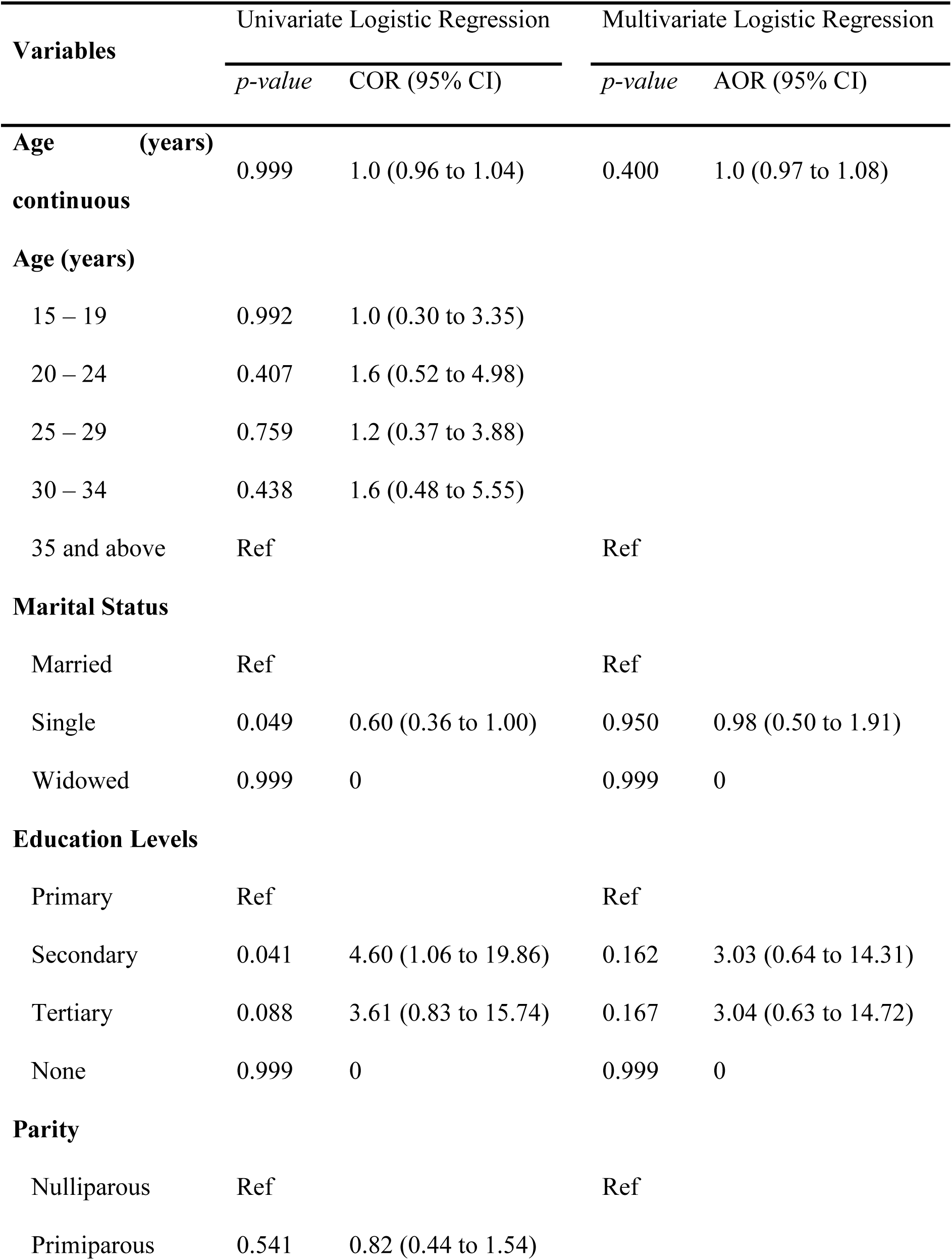
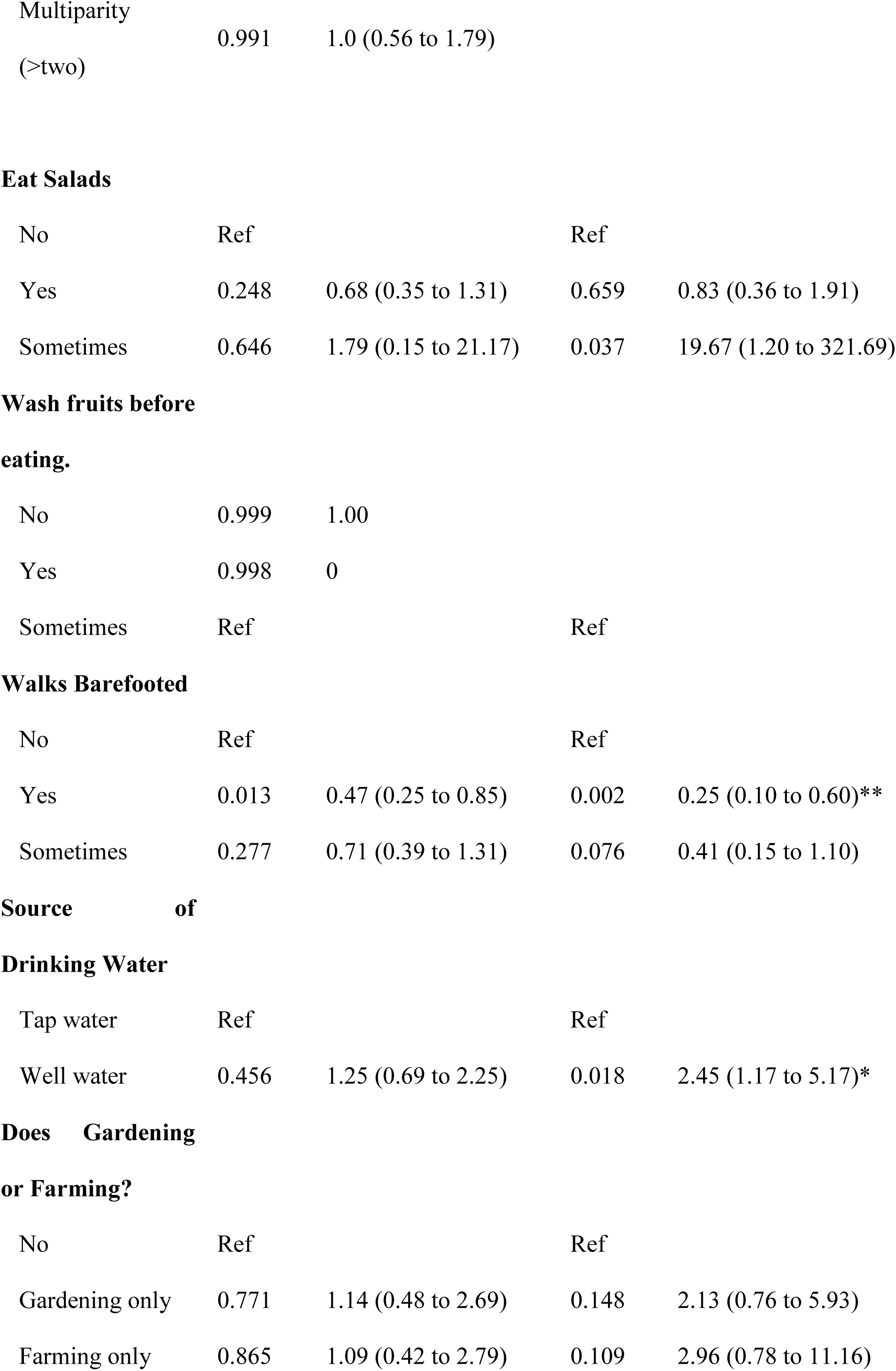
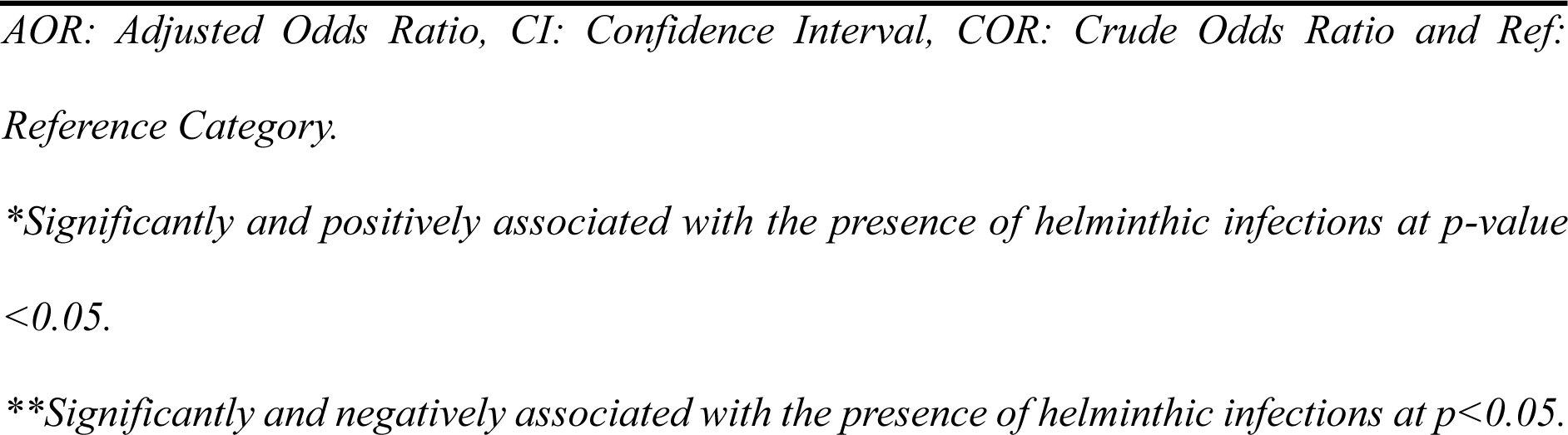
Assessment of risk factors associated with intestinal helminths among pregnant women attending ANC in Kabwe, Zambia from November 2021 to February 2022.

**Table 6:** Chi-square Test of Association between the Levels of Haemoglobin and Results of Stool Microscopy for Parasites among pregnant women attending ANC in Kabwe, Zambia from November 2021 to February 2022.

| Variables | Haemoglobin Level g/dl |  | Total | p-value |
| --- | --- | --- | --- | --- |
|  | ≥11 g/dl<br>(n=287) | <11 g/dl<br>(n=155) |  |  |
| **Microscopy Results for Parasites** |  |  |  | <0.0001 |
| No Parasites | 264 (92%) | 103 (66.5%) | 367 |  |
| Parasites Present | 23 (8%) | 52 (33.5%) | 75 |  |
| **Parasites** |  |  |  | <0.0001 |
| *Trichuris trichiura* | 0 | 1 (0.6%) | 1 |  |
| *Hookworms and S. stercolaris* | 0 (0%) | 2 (1.3%) | 2 |  |
| *Hookworms and Ascaris lumbricoides* | 0 (0%) | 3 (1.9%) | 3 |  |
| *Ascaris lumbricoides only* | 3 (1%) | 7 (4.5%) | 10 |  |
| *Hookworms only* | 20 (7%) | 39 (25.2%) | 59 |  |

### Univariate and Multivariate Logistic Regression Results for Factors Associated with Parasitic Infections in Pregnant Women attending ANC in Kabwe, Zambia from November 2021 to February 2022

The age of the women was not significantly associated with the risk of parasitic infections, as indicated by the p-value of 0.999 and the odds ratio (OR) of 1.0 (95% CI: 0.96 to 1.04) in both the univariate and multivariate analyses. Marital status was also non-significantly associated with the risk of parasitic infections.

Women with higher levels of education were more likely to be infected with parasitic infections. Specifically, women with secondary education (OR: 4.60, 95% CI: 1.06 to 19.86) and tertiary education (OR: 3.61, 95% CI: 0.83 to 15.74) were more likely to be infected compared to those with primary education or no education. The parity of the women was not significantly associated with the risk of parasitic infections in primiparous women (OR: 0.82, 95% CI: 0.44 to 1.54) and multiparous women (>two) (OR: 1.00, 95% CI: 0.56 to 1.79).

Eating salads was associated with a higher risk of parasitic infections (p-value = .037, OR: 19.67, 95% CI: 1.20 to 321.69), Walking barefooted was found to be significantly associated with an increased risk of parasitic infections (p-value = .076, OR: 0.25, 95% CI: 0.10 to 0.60)

The source of drinking water was found to be significantly associated with the risk of parasitic infections, with women who used well water having a higher odds ratio (p-value = .018, OR: 2.45, 95% CI: 1.17 to 5.17) compared to those who used tap water. Lastly, the analysis showed that women who engaged in gardening or farming only had a higher odds ratio (OR: 2.13, 95% CI: 0.76 to 5.93) compared to those who did not engage in these activities.

### Association between the Levels of Haemoglobin and Results of Stool Microscopy for Parasites

The results of the chi-square test of association between the levels of haemoglobin and the results of stool microscopy for parasites among the 442 study participants in Table 7 reveal a strong statistical association between the two variables. The p-value of <0.0001 indicates that the observed association is highly unlikely to occur by chance, suggesting that there is a real association between haemoglobin levels and the presence of parasites. The results show that among pregnant women with haemoglobin levels above 11g/dl, 92% had no parasites detected in their stool samples, while only 8% had parasites present. In contrast, among pregnant women with haemoglobin levels below 11g/dl, only 66.5% had no parasites detected, while 33.5% had parasites present. This suggests that pregnant women with lower haemoglobin levels are more likely to have intestinal helminth infections.

The specific types of parasites detected also vary significantly by haemoglobin level.

Table 7 shows the presence of hookworms and *Ascaris lumbricoides* is associated with lower haemoglobin levels in pregnant women there was a significant difference in the distribution and median haemoglobin levels between pregnant women with intestinal helminthic infections and those without.

### Mann-Whitney U Test for Participants with Worm Infestation

**Table 7:**
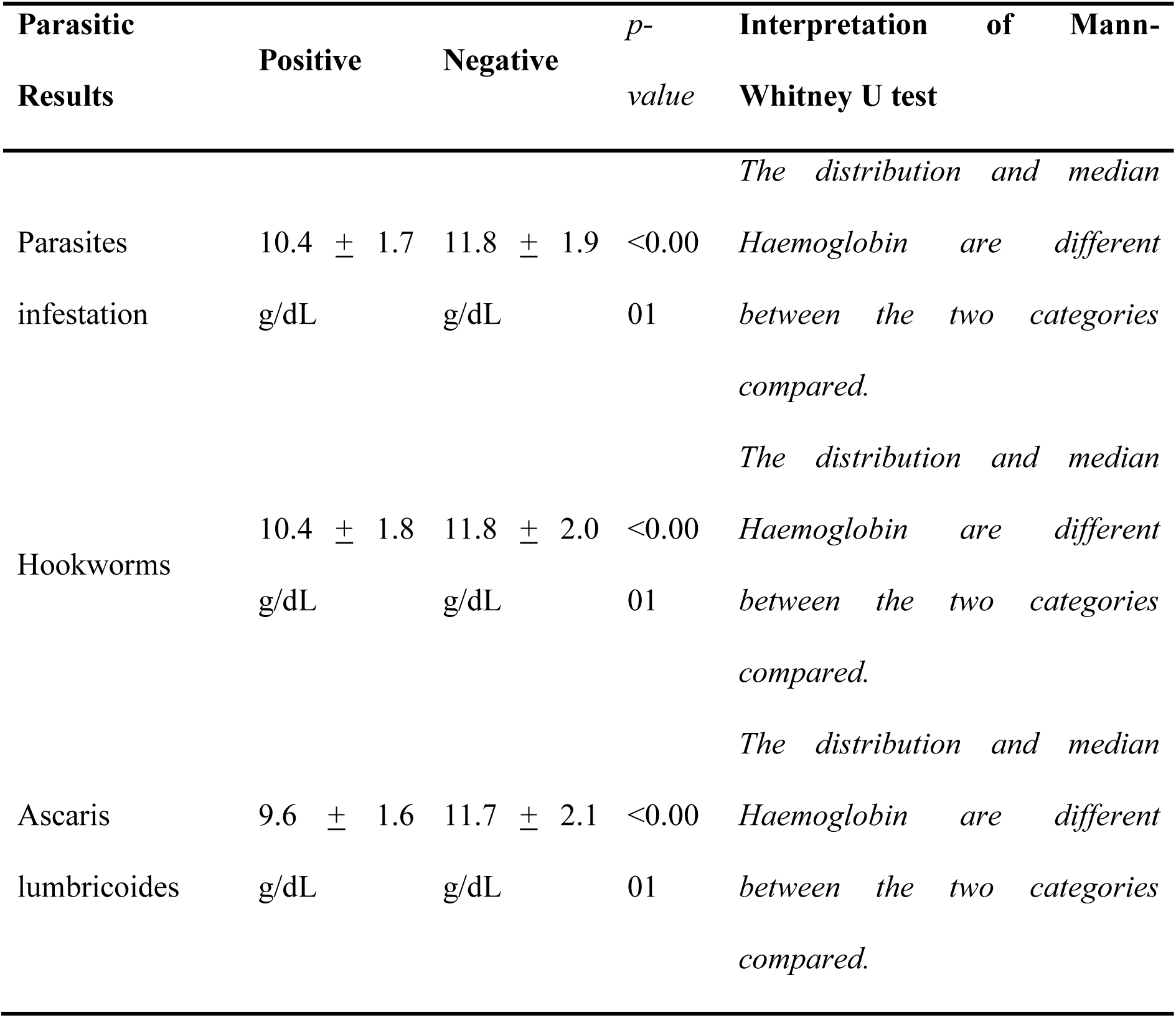
Results of Mann-Whitney U Test for Participants with worm infestation.

## Discussion

Zambia is a developing country where IPIs are major public health problems. Previous studies carried out in Zambia recorded a high prevalence of IPIs [3, 4, 5]. The burden of intestinal parasites, particularly the soil-transmitted helminths (STHs), is often very high in school children [4, 5].

The prevalence of intestinal helminths in pregnant women in this study (17%) is similar to those reported among Zambian children in Kafue district 17.9% [5] and comparative with those reported in Chililabombwe district (14.1%) [4].The results are also similar to those reported among pregnant women in South East Nigeria (18.6%) [10]. It is again lower than data from South West Ethiopia (41%) [11] and Southern Nigeria (48.3%) [12]. Conversely, prevalence is higher than that reported in Kenya (13.8%) [13] and Ethiopia (13.7%) [14]. This difference could be due to geographic variations, lack of sensitivity to diagnostic methods of the different intestinal helminths, differences in socio-economic conditions of the study populations and individual behavioural habits, sanitation, presence of water, and environmental conditions such as humidity, temperature, and awareness levels of pregnant women about ANC.

The predominant intestinal helminth in this study was Hookworm (13.3%), *Ascaris lumbricoides* (2.3%) *Trichuris trichuira* (0.2%) and *S. stercoralis* (0.5%). These findings are consistent with those of studies conducted in Nigeria 14.6% [15] and Ethiopia [11, 16, 17] where authors reported that Hookworm was the predominant intestinal helminth. Conversely, the predominance of Hookworm in this study is different from a study conducted in Kenya [13] where *Ascaris lumbricoides* was found to be the predominant species. The possible difference could be due to the geographic variation, the tropical nature of our study area which favours Hookworm transmission and the difference in the habit of walking barefoot which is more practised in Zambia specifically in the study site, when the people are at home. The low prevalence of *Trichuris trichura* and *S. stercoralis* infections in this study is consistent with those of a study in Ethiopia [16] and Tanzania [18].

The high prevalence of parasitic findings in this study suggests that these parasites are common in the study population and may contribute to adverse pregnancy outcomes such as iron deficiency anaemia, malnutrition and impaired foetal growth. [19, 20] making it important to consider these infections in the management of pregnant women.

Anaemia in pregnancy should be prevented and strictly managed as recommended by the World Health Organization [21]. The total prevalence of anaemia in this study was 35.1%. This figure is comparable to previous similar studies in Western Ethiopia 36.6% and Cameroon 33.5% [22].On the other hand, this prevalence is lower than that reported in Malawi (57.1%)[23] and Peru (50%)[24]. This difference could be explained by the comparative oldness of the previous data, the inability to use sensitive techniques to determine haemoglobin level in the current study, the difference in altitude, better awareness of pregnant women in including diversified food diets and also prevention of intestinal helminths.

In this study, anaemia was significantly associated with intestinal helminthic infection specifically with Hookworms because Hookworms ingest blood and damage the intestinal wall causing bleeding thus contributing to anaemia by causing blood loss directly through ingestion and mechanical damage of the mucosa, and indirectly, by affecting the supply of nutrients necessary for erythropoiesis thus accounting for the highest prevalence of anaemia in hookworm infested participants. Anaemia can be caused by several factors, which were not investigated in this study. Other probable causes of anaemia include; iron deficiency, heavy menstrual bleeding in adolescent girls and women, problems in red blood cell production, inherited disorders (thalassemia or sickle cell anaemia), infections such as malaria, toxins like alcohol that damage the bone marrow, malabsorption of nutrients in the diet, chronic inflammatory diseases that reduce the use of iron in the body and bleeding during a surgery [25] Symptoms of anaemia are tiredness, dizziness, weakness and shortness of breath, among others. The WHO estimates that 37% of pregnant women globally are anaemic. Therefore, anaemia is a serious global public health problem, especially in pregnant women [26].

Walking barefoot, drinking well water and eating uncooked vegetables (salads) were independent predictors of intestinal helminthic infection in the present study. These findings are comparable with the studies in Ethiopia [16, 17] found that walking barefooted had a positive association with intestinal helminths and was associated with an increased risk of hookworm infection. On the other hand, data from Chililabombwe, Zambia [4] reported that residence, household income and overcrowding were significant factors for intestinal helminthic infection in children. This difference might be due to differences in geographic locations and behavioural factors of the different study participants. Most rural and peri-urban pregnant women walk barefoot when they are at home; even those women who have shoes do not wear them regularly. Walking barefoot may predispose to hookworm infection because the infective stage of the parasite develops in soil.

In addition, the results also showed that using well water was significantly associated with an increased risk of parasitic infections, which is consistent with previous studies in Kenya [27]. One possible risk factor for STH infection is a household water supply. According to studies conducted by [28, 29, 30].

The findings also showed that eating salads was associated with a higher risk of parasitic infections, which is consistent with previous studies that have found that eating raw or undercooked food can increase the risk of parasitic infections [31].

Furthermore, the study’s findings also highlight significant proportion of women (64% and 25.8%) engage in gardening and farming activities, respectively, which can increase their exposure to contaminated soil increasing the risk of intestinal helminthic infections among pregnant women . This is consistent with a study in Kenya [32].

Finally, the study observed that a significant proportion of women (87.3%) had not dewormed in the past six months or more suggesting that there is a low level of awareness and practice of deworming among healthcare providers and pregnant women in this region. Deworming is an effective strategy for preventing intestinal helminthic infections among pregnant women [33]. Therefore, increasing access to deworming services may be an effective strategy for reducing the risk of intestinal helminthic infections among pregnant women in this region.

The findings of this study revealed a strong statistical association between the levels of haemoglobin and the results of stool microscopy indicating that pregnant women with lower haemoglobin levels are more likely to have intestinal helminth infections. This is consistent with the findings of a study conducted in Nepal [34] and Ghana [35]. Hookworms were found in 25.2% of women with haemoglobin levels below 11g/dL. *Ascaris lumbricoides* was found in 4.5% of women with haemoglobin levels below 11g/dL. These findings are consistent with those of a study conducted in Indonesia [36].The findings of this study suggest that the presence of intestinal helminthic infections is a significant risk factor for anaemia in pregnant women (p<0.0001). This is consistent with the findings of other studies [37, 38, 39] that have reported a strong association between helminthic infection and anaemia.

The study was facility-based therefore, it is not possible to generalise the findings to all pregnant women left in the community and did not attend antenatal care services. In this study, we used formol-ether concentration technique for the detection of intestinal helminths. We had not used other floatation techniques and other sensitive methods which might result in underestimation of the prevalence of intestinal helminths. Further, we had not used automated machines to measure haemoglobin levels which may again render the prevalence of anaemia in this study inaccurate. Having these limitations, we believe that the results of this study can contribute to policymakers and clinicians to prevent helminths and anaemia during pregnancy.

## Conclusion

The study ascertained that Soil-Transmitted Helminths are prevalent among pregnant women in the study area. The prevalence of intestinal helminths in the study area was significantly high. Four different species of intestinal helminths were identified with a predominance of soil-transmitted helminths (Hookworm and *Ascaris lumbricoides*). Walking barefoot, consuming raw vegetables(salads) and drinking water from the well were identified as independent risk factors that significantly increase intestinal helminths in pregnant women with p-value <0.05. The prevalence of anaemia was significantly high among pregnant women infected with intestinal helminths specifically with Hookworms. Public health measures, regular ANC follow-up with an intensive diagnosis of intestinal helminths and further longitudinal studies are recommended.

## Data Availability

The datasets used during the study are available from the corresponding author can be accessed on resonable request

## Acknowledgements

We would like to thank the University of Zambia Management and staff for the academic and administrative support, all study participants for their cooperation, and the administrative staff of respective health centres for all the support given to carry out this study. Moreover, we would like to thank the data collectors and study participants without whom the research would not be a reality.

## Consent for publication

Not applicable.

## Availability of data and materials

The datasets used and/or analysed during the current study are available from the corresponding author on reasonable request.

## Funding

This work was not a funded project

## Conflicts of interest

Financial interests: The authors declare that they have no relevant financial or non-financial interests to disclose and they have no conflicts of interest to declare that are relevant to the content of this manuscript.

## Authors’ contributions

MK and NMS-M designed the study. MK conducted sample collection. AS analysed the data. MK and NMS-M interpreted the data. WH and MK drafted the manuscript, NMS-M edited and reviewed the manuscript.NMS-M supervised the implementation of the project. All authors read and approved the final manuscript.

## Notes

### Competing Interest Statement

The authors have declared no competing interest.

### Funding Statement

The author(s) received no specific funding for this work.

### Author Declarations

The University of Zambia Health Sciences Research Ethics Committee Protocol ID 202112030113.

